# Tobacco smoking and vaping in pregnancy and neonatal outcomes: results from a UK cohort study

**DOI:** 10.1101/2025.11.03.25339380

**Authors:** Anthony A Laverty, Oluwaseun Esan, Ian Sinha, Parris J Williams, Filippos T Filippidis, Nicholas S Hopkinson

## Abstract

**Background:** Although the harms of smoking during pregnancy are well established, there is less evidence on the impacts of vaping.

**Methods:** We analysed data on caregiver-reported birthweight, low birthweight (LBW, <2.5kg), postnatal hospital stay, and admission to a neonatal care unit from interviews for 1,896 babies in the UK Early Life Cohort Feasibility Study (9–10 months sweep). Smoking and vaping during pregnancy were classified as “yes” vs “never” to form four mutually exclusive exposure groups. Models adjusted for sociodemographic factors and survey weights. Linear, logistic, and negative binomial regressions were used. Sensitivity analyses included exposure frequency categories and models using logged birthweight or household income.

**Findings:** Most mothers (90.1%, N=1528) neither smoked nor vaped; 2.6% (N=44) vaped only, 5.5% (N=93) smoked only, and 1.8% (N=31) were dual users. Mean birthweights were 3.31 kg where mothers neither smoked nor vaped and 3.00kg where mothers smoked but did not vape. Compared to unexposed mothers, adjusted models showed lower birthweight for babies of mothers who smoked (-0.34kg, p<0.001) and where mothers were dual users (-0.39kg, p=0.001).Smoking exposure demonstrated a dose-response association with lower birthweight (trend p=0.001) and LBW (p<0.001), while vaping showed no associations. Dual use was associated with longer hospital stay (+0.88 days; p=0.001); smoking or vaping alone were not. No group differed in the incidence of neonatal admission.

**Interpretation:** Results underline the importance of quitting smoking completely during pregnancy. Although pregnant women should be advised to quit vaping, this must not be at the expense of reverting to smoking.

**Funding:** NIHR School for Public Health Research

## INTRODUCTION

The harms of tobacco smoking in pregnancy including stillbirth, miscarriage, and pre-term birth are long established [1–4]. More recently, the possibility that use of e-cigarettes in pregnancy may also have adverse effects has given rise to concern. Because e-cigarettes can provide an alternative source of nicotine to combustible tobacco, many people have used them to quit smoking [5,6]. However, failure to regulate the marketing and sales of these devices [7–9] has led to widespread uptake among young people [10], including many who have never smoked [6]. This may translate into vaping during pregnancy, especially considering that risk perceptions and advice to quit by health professionals may differ for smoking and vaping. Some people who vape during pregnancy will also have smoked previously, and some will be dual users.

Evidence from biomarkers of exposure and harm [11] indicate that, because of the absence of combustion, vaping is likely to be substantially less harmful overall than smoking, though the precise quantification of this difference for individuals over potentially decades of use remains to be established. Both smoking and vaping behaviours are largely driven by nicotine addiction, and circulating nicotine levels tend to be similar or lower in people who vape compared to people who smoke [11]. There is limited data so far specifically in pregnancy, and a recent UK Office for Health Improvement & Disparities report notes that the “effects of vaping on fetal development and pregnancy outcomes remain in particular need of research” [11].

Globally, there is limited data on the precise prevalence of maternal smoking during pregnancy, although systematic review evidence from 2018 finds a global prevalence of 1.7% with substantial country variation [10]. Other reviews have highlighted that while there have been declines in smoking during pregnancy, social inequalities are strongly related to this behaviour, and that these inequalities are persisting in many countries [12]. There is understandably less data on the prevalence of vaping during pregnancy, with a 2021 review finding insufficient data to estimate prevalence. This did however, note that in 12/14 studies, women reported using e-cigarettes to quit smoking tobacco during pregnancy [13]. A 2024 review of potential health harms of vaping in pregnancy concluded that the quality of the existing evidence on this issue limited definitive conclusions [14]. Limitations included small sample sizes in many studies, lack of comparison of exclusive smoking to exclusive vaping, and a lack of information in many studies on factors such as maternal age, ethnicity and socio-economic status.

There is a need therefore for data to study the effects of vaping on pregnancy outcomes. Birth cohorts, with prospective collection of data, provide an opportunity to address this evidence gap.

## METHODS

The UK Early Life Cohort Feasibility Study: Age 9-10 months sweep used computer assisted face-to-face interviews between September 2023 and September 2024 to collect data from the caregivers of 1976 babies [15]. The Early Life Cohort Feasibility study was known to participants as Generation New Era and recruited a representative birth cohort of babies under 1 year old across all four nations of the UK [16]. It collected data on families using trained interviewers with up to four interviews per baby covering both main caregivers as well as secondary caregivers. National birth records were used to inform the sampling frame for representativeness, and 49% of invited families agreed to take part in the study. Sample boosts were undertaken in specific populations and groups; In England and Wales, 40% of families recruited lived in the most deprived fifth of England, over half of babies were not White British and close to 15% of mothers were under the age of 25 years.

Our primary outcome was birthweight in kilograms (kg) as reported by caregivers. We also included three secondary outcomes: being born at a Low Birth Weight (LBW) of under 2,500 grams, length of hospital stay (in days) and admission to a neonatal care unit (categorised as yes vs. no). For length of stay and admission to a neonatal care unit outcome, which were caregiver-reported, we excluded home births and babies still in hospital (n=55).

To assess tobacco smoking mothers were asked: ‘During your pregnancy with your baby did you smoke cigarettes at all?’ We categorised this as no (‘No’) and ever (‘Yes, occasionally’;’ Yes, most days’ and ‘Yes, every day’). To assess vaping mothers were asked: ‘During your pregnancy with your baby did you use e-cigarettes/vapes or waterpipes products at all?’ We categorised this as no (‘No’) and ever (‘Yes, occasionally’; ‘Yes, most days’ and ‘Yes, every day’). This allowed a four-way exclusive categorisation of never smoking nor vaping during pregnancy; vaping but not smoking during pregnancy; smoking but not vaping during pregnancy; and dual use of both products during pregnancy.

We also included data on sex of babies, UK nation (England, Scotland, Wales, Northern Ireland), Index of Multiple Deprivation in five groups, maternal age (≤ 29 years vs. ≥ 30 years), maternal ethnic group (White, Asian or Asian British, Black or Black British, Mixed and Other Ethnic groups).

### Missing data

The original sample size was 1,973 from which we excluded data on 82 twins. For the remaining 1,891 births 117 (6·2% of sample) were missing the primary outcome of birthweight and 195 (10·3% of sample) were missing the primary exposure variable of smoking and vaping status. Levels of missingness across our covariates was low: baby sex and country had no missing data, IMD had 1 missing value, maternal age had 3 missing values, and maternal ethnicity had 5 missing values. As overall levels of missingness were low, we did not impute missing data. There are therefore different numbers included in different regression results which we note in relevant tables.

### Analyses

Analyses were guided by a directed acyclic graph (DAG) we developed to represent relationships between the key factors in our analysis (**Figure 1**). Our primary analyses used linear regression to compare birthweight across these four mutually exclusive smoking and vaping groups, controlling for factors listed above. We used logistic regression with the same covariates for analyses of LBW and admission to a neonatal care unit as outcomes. For analyses of the number of days in hospital post-partum, we used negative binomial regression as the dispersion parameter was significantly greater than 0 (p<0.001). All regression models also adjusted for survey weights provided by the data collectors to control for the complex survey design and differential non-response.

**Figure 1:**
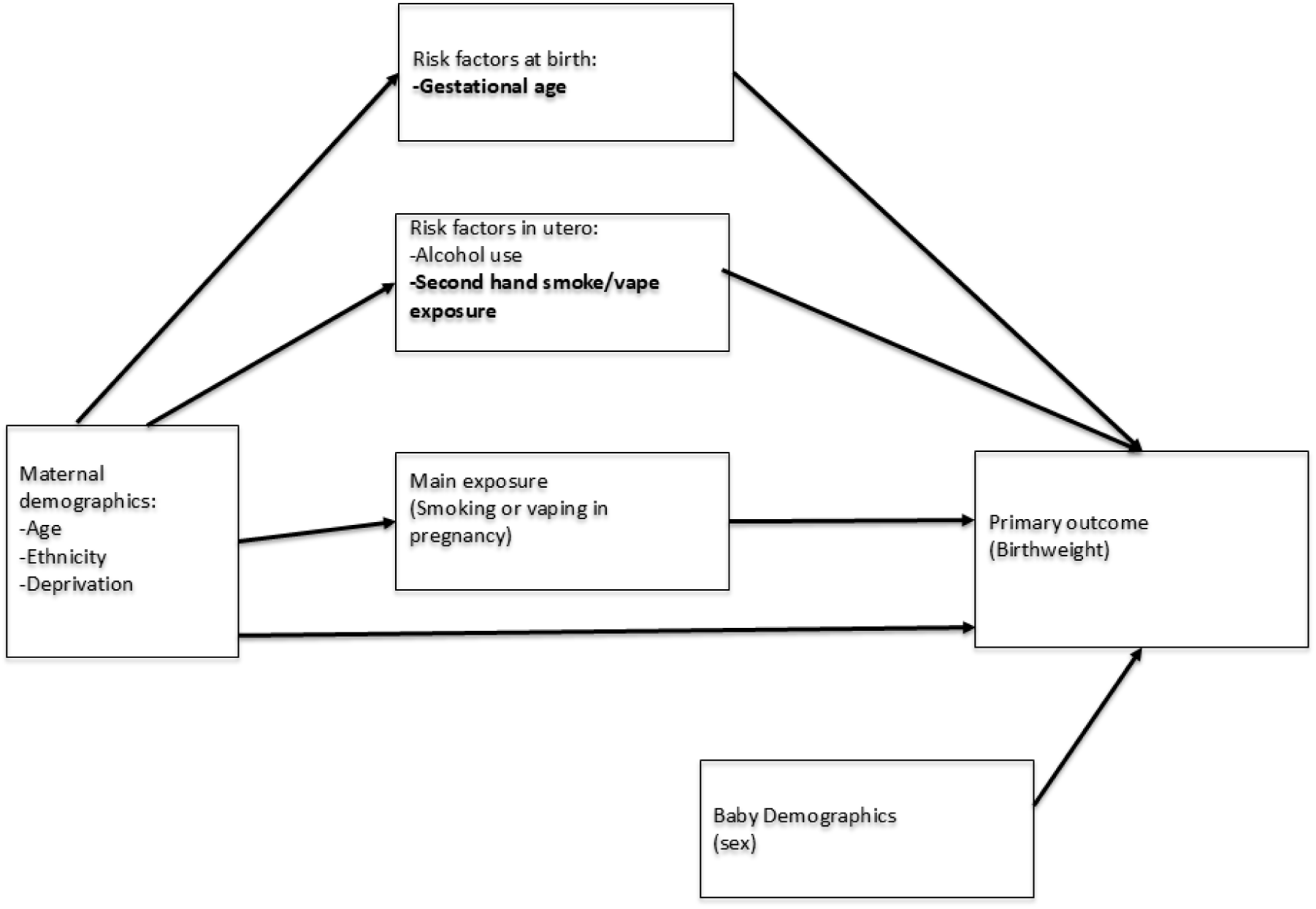
Directed Acyclic Graph (DAG) depicting the relationship between smoking, vaping and birthweight Unmeasured factors are noted in bold.

Although the sample size was fixed for this analysis of birth cohort data, sample size calculations based on the numbers in each exposure group and the means in these groups estimated an 83·4% power to detect a difference of 300g difference between mothers who vaped and non-users. Corresponding values for this difference between non-users and mothers who smoked and dual users were 98·7% and 72·1% respectively.

For our primary outcome of continuous birthweight there was some evidence of right-skew in model residuals. For this reason, we also include linear regression analyses on the logged outcome as a supplementary analysis. Analyses were conducted in Stata 15·1.

### Sensitivity analyses

We performed multiple sensitivity analyses to examine robustness of our main findings. While our main analyses use ever smoking or vaping as the primary exposure, in sensitivity analyses we used a more granular assessment of smoking and vaping in pregnancy. These groups were: no smoking during pregnancy; occasional smoking during pregnancy; smoking most days during pregnancy; and smoking every day during pregnancy. We used the same frequency categories for vaping. For both of these analyse, we used the same models and structures as above, and did not adjust for vaping in the models of smoking frequency (and vice versa). In these analyses we also present a test for linear trend to assess dose response across frequency categories.

Due to some evidence of non-linearity of our primary outcomes, we conducted analyses which had logged birthweight, and report findings as percentage differences. As IMD is an imperfect measure of deprivation, we performed analyses using household income as a covariate. This was collected in four groups: less than £330 a week; £330-£530 a week; £530-£800 a week; and more than £800 a week.

## RESULTS

Most mothers (90·1%, N=1,528) neither smoked nor vaped during pregnancy (**Table 1**). 2·6% (N=44) of the sample vaped but did not smoke, 5·5% (N=93) smoked but did not vape and 1·8% (N=31) were dual users during pregnancy.

**Table 1:** Birth outcomes by maternal smoking and vaping status.

|  | **Neither**<br>**smoking nor**<br>**vaping**<br>**(N=1528)** | **At least**<br>**occasional**<br>**vaping, no**<br>**smoking**<br>**(N=44)** | **At least**<br>**occasional**<br>**smoking, no**<br>**vaping (N=93)** | **At least**<br>**occasional**<br>**dual use**<br>**(N=31)** |
| --- | --- | --- | --- | --- |
| % of sample | 90.1 | 2.6 | 5.5 | 1.8 |
| Mean birthweight (kg) | 3.31 | 3.38 | 3.00 | 3.06 |
| SD | 0.65 | 0.67 | 0.58 | 0.77 |
| Days in hospital (mean) | 2.36 | 2.05 | 2.23 | 3.76 |
| SD | 2.46 | 1.95 | 2.24 | 4.68 |
| Neonatal unit stay (n) | 187 | 4 | 13 | 4 |
| % | 12.3 | 9.1 | 14.0 | 13.3 |
| LBW (n) | 122 | 2 | 20 | 5 |
| % | 8.3 | 4.6 | 21.5 | 17.2 |

Mean birthweights were 3·31kg (SD 0·65) for babies whose mothers neither smoked nor vaped. Birthweights were 3·38kg (SD 0·67) where mothers vaped but did not smoke; 3·00kg (0·58) where mothers smoked but did not vape; and 3·06kg (SD 0·77) where mothers both smoked and vaped. Mean days in hospital ranged from 2·05 (SD 2·46) where mothers vaped but did not smoke to 3·76 (SD 4·68) among dual users. The percentage of babies admitted to a neonatal unit during their stay ranged from 9·1% where mothers vaped but did not smoke, to 14·0% for those who smoked but did not vape. The percentage of babies born LBW was 8.3% where mothers neither smoked nor vaped and 17.2% where mothers were dual users during pregnancy.

Results from fully adjusted regression models found no statistically significant difference in birthweights between mothers who vaped and mothers who neither smoked nor vaped (+0·00kg, 95%CI -0·14 to 0·15kg) (**Table 2**). Birthweights were lower for babies of mothers who smoked (-0·34kg, 95%CI -0·46 to -0·22kg) and where mothers were dual users (-0·39kg, 95%CI -0·62 to -0·17kg). Female babies had lower birthweights than male babies (-0·12kg, 95% CI -0·19 to -0·17kg), while babies born to Asian or Asian British mothers had lower birthweights than babies born to White mothers (-0·46kg, 95%CI -0·59 to -0·32kg), as did babies born to Black or Black British mothers (-0·33kg, 95%CI -0·47 to -0·19).

**Table 2:** Results from linear regression model of maternal smoking/vaping during pregnancy and birthweight (kg)

| N=1,627 | Coefficient (%) | p value | Lower CI | Upper CI |
| --- | --- | --- | --- | --- |
| Neither smoking nor vaping | ref | ref | ref | ref |
| At least occasional vaping, no smoking | 0.00 | 0.962 | -0.14 | 0.15 |
| At least occasional smoking, no vaping | -0.34 | <0.001 | -0.46 | -0.22 |
| At least occasional both | -0.39 | 0.001 | -0.62 | -0.17 |
| Male babies | ref | Ref | ref | ref |
| Female babies | -0.12 | 0.001 | -0.19 | -0.05 |
| England | ref | Ref | ref | ref |
| Northern Ireland | 0.15 | 0.002 | 0.06 | 0.24 |
| Scotland | 0.00 | 0.978 | -0.09 | 0.09 |
| Wales | 0.04 | 0.342 | -0.04 | 0.12 |
| Most deprived areas | ref | ref | ref | ref |
| 1 | -0.01 | 0.771 | -0.10 | 0.08 |
| 2 | 0.05 | 0.352 | -0.06 | 0.16 |
| 3 | 0.01 | 0.859 | -0.10 | 0.12 |
| Least deprived | 0.10 | 0.042 | 0.00 | 0.20 |
| Maternal age<30 | ref | ref | ref | ref |
| Maternal age30+ | 0.00 | 0.93 | -0.07 | 0.06 |
| Maternal ethnicity: White | ref | ref | ref | ref |
| Asian or Asian British | -0.46 | <0.001 | -0.59 | -0.32 |
| Black or Black British | -0.33 | <0.001 | -0.47 | -0.19 |
| Mixed and Other | -0.21 | 0.100 | -0.46 | 0.04 |

Results from fully adjusted model controlled for all covariates listed here
CI = Confidence Interval
ref = reference category

Adjusted logistic regression models found that, compared to where mothers neither smoked nor vaped, that babies born to mothers who smoked but not vaped were more likely to be LBW (odds ratio 3·96, 95%CI 2·22 to 7·08) (**Table 3**). Higher likelihood of LBW was also observed for babies where mothers were dual users compared to where mothers neither smoked nor vaped (odds ratio 3·67, 95%CI 1·24 to 10·83). The adjusted negative binomial models found a greater number of days in hospital post-partum for dual users (+0·88 days, 95%CI 0·39 to 1·38 days) but no statistically significant associations of smoking or vaping alone. The logistic regression model did not identify any differences in likelihood of being admitted to a neonatal care unit across categories of smoking/vaping.

**Table 3:** Results from regression models of maternal smoking/vaping during pregnancy and Low Birth Weight (<2,500g, logistic regression), days in hospital (negative binomial regression) and being cared for on neonatal unit (logistic regression)

|  | Coefficient | p-value | Lower CI | Upper CI |
| --- | --- | --- | --- | --- |
| Low Birth Weight (presented as odds ratios from logistic regression) |  |  |  |  |
| N=1,627 |  |  |  |  |
| Neither smoking nor vaping | ref | ref | ref | ref |
| At least occasional vaping, no smoking | 0.54 | 0.447 | 0.11 | 2.66 |
| At least occasional smoking, no vaping | 3.96 | <0.001 | 2.22 | 7.08 |
| At least occasional both | 3.67 | 0.019 | 1.24 | 10.83 |
| Days in hospital (presented as days from negative binomial regression models) |  |  |  |  |
| N=1,582 |  |  |  |  |
| Neither smoking nor vaping | ref | ref | ref | ref |
| At least occasional vaping, no smoking | -0.07 | 0.602 | -0.35 | 0.20 |
| At least occasional smoking, no vaping | -0.02 | 0.842 | -0.25 | 0.20 |
| At least occasional both | 0.88 | 0.001 | 0.39 | 1.38 |
| Being admitted to neonatal unit (presented as odds ratios from logistic regression) |  |  |  |  |
| N = 1,680 |  |  |  |  |
| Neither smoking nor vaping | ref | ref | ref | ref |
| At least occasional vaping, no smoking | 0.65 | 0.462 | 0.21 | 2.04 |
| At least occasional smoking, no vaping | 1.02 | 0.943 | 0.53 | 1.96 |
| At least occasional both | 1.42 | 0.535 | 0.47 | 4.31 |

Analyses of the frequency of maternal smoking during pregnancy and outcomes are shown in **Appendix Table 1**. These find evidence of a dose response relationship between frequency of maternal smoking and birthweights (p-value for linear trend = 0·001). For example, babies born to mothers who smoked occasionally during pregnancy were -0·35kg smaller (95%CI -0·49 to -0·20kg) than babies born to mothers who did not smoke in pregnancy. Logistic regression analysis of LBW found that all levels of maternal smoking during pregnancy were associated with increased likelihood of babies being born LBW and that there was evidence of dose response here (p value for linear trend <0.001). For example, babies born to mothers who smoked occasionally were over three times (odds ratio 3·18, 95%CI 1.32 to 7·68) more likely to be LBW. We did not identify differences in days spent in hospital nor admission to a neonatal care unit likelihood by maternal smoking frequency.

Analyses using frequency of vaping during pregnancy did not identify a statistically significant trend for birthweights (p value for linear trend 0·507), although babies born to mothers who vaped occasionally were lighter than those born to non-vaping mothers (-0·22kg, 95%CI -0.42 to -0.01kg) **(Appendix Table 2).** There was similarly limited evidence of dose response across LBW, days spent in hospital and likelihood of admission to a neonatal care unit. Sensitivity analyses using logged birthweight as an outcome gave similar results as did analyses using income rather than IMD as a covariate (**Appendix tables 3 and 4**).

## DISCUSSION

The main finding from this analysis of data from a cohort study of children born in 2022/23 is that while smoking during pregnancy was associated with reduced birthweight and likelihood of being born at a low birth weight. These associations were not seen with vaping.

There was no statistically significant association of smoking or vaping alone with days spent in hospital post-partum or with admission to the neonatal care unit.

### Significance of findings

While birthweight is a useful overall measure of development, including lung development during pregnancy [17] the absence of an effect of vaping on birthweight should not be taken to indicate that no risk exists. Although many of the toxic components of cigarette smoke are either absent or present at much lower levels, e-cigarette vapour contains nicotine and a variety of chemicals produced when flavours are heated [11]. It may also contain trace quantities of metals derived from the heating element. The optimum situation is for mother and foetus to be exposed to neither cigarette smoke nor e-cigarette vapour. However, our results provide useful information about the relative risks of smoking and vaping in pregnancy.

This analysis was conducted in the UK, where regulations limit the nicotine content of e-cigarettes to 20mg/ml, similar to most European countries. Other countries, notably the United States of America, have no legal limits of nicotine content and vaping products typically contain much higher levels of nicotine [18]. As such, our findings may not be generalisable to settings where the composition of vaping products is substantially different.

Nicotine replacement therapy is licensed for use during pregnancy, with the most recent Cochrane review identifying no differences in adverse outcomes between NRT treatment in this context and control groups [19]. Clinical trial data suggest that use of e-cigarettes in pregnancy leads to a reduced incidence of LBW (9·8% vs 14·8%) compared to NRT [20]. This difference in LBW incidence may in part be due to e-cigarette use being a more effective smoking cessation agent than NRT [5].

### Methodological issues

There are limitations in this study to consider. As it was based on a pre-existing birth cohort with a fixed sample size there were limitations to the power to detect group differences, especially in relation to maternal vaping and maternal dual use, as both of these behaviours are relatively rare. Nonetheless, the analyses did detect differences between maternal dual users and non-users in our primary analyses, while estimates for the difference between vaping and non-users was very close to zero.

Our analyses were guided by the development of the DAG to depict key relationships, which highlights potentially important missing data here. We were unable to conduct formal mediation analysis and were unable to control for some potential mediators due to missing data. Thus, while models were adjusted for factors including deprivation and maternal age, there may be residual confounding from other individual characteristics related to smoking or vaping behaviour. We also did not include data on either second-hand smoke or second-hand vaping exposure in utero. While this is a limitation, impacts of second-hand exposure compared to smoking and vaping are likely smaller than for the direct effects of these behaviours. We also did not have data on gestational age which would allow for a more accurate characterisation of outcomes, including whether the issue is fetal growth restriction or being small for gestational age [21,22]. Nonetheless, we did control for a range of socio-demographic factors which limits the importance of this omission [23].

The results are based on self-report rather than objective measures of smoke or nicotine exposure. The question about vaping included the use of waterpipe, but we disregarded this in our analysis due to the very low prevalence of waterpipe use in this population[24] and given that this is a form of combusted tobacco it would anyway have tended to increase any apparent detrimental effects in this group. Finally, these findings demonstrate the importance of collecting such birth cohort data to obtain data not routinely collected in healthcare data. They also highlight the need for data linkage between such cohort data and other data sources which would allow assessment of key health indicators. Such data linkage as well as analyses with larger sample sizes will be needed to make any assessment of infrequent adverse events.

## Conclusion

These results underline the importance of quitting smoking completely to improve pregnancy outcomes. Although vaping did not appear to have an impact on birthweight, the precautionary principle means that people who vape and who are pregnant or planning to become pregnant should be advised to quit vaping, though not at the expense of going back to smoking which is substantially more harmful. In addition, where an individual has quit smoking by switching to vaping the importance of avoiding relapse to smoking and of avoiding dual use should be highlighted.

## Data Availability

All data is available from the UK Data Archive here https://ukdataservice.ac.uk/

## Statements

### Contributorship

AL and NH conceived of the idea for the study. All authors designed the study and interpreted analyses and findings. AL and NHS wrote the first draft which all authors edited, and all authors approved the final version. PW and AL have both accessed and verified the underlying data and AL is guarantor.

### Data Sharing

Data for this study are publicly available from the UKDataService

### Ethics approval

This secondary analysis of anonymised data did not require ethical approval.

### Data sharing

Data available from UK Data Service https://ukdataservice.ac.uk

---

**Appendix table 1:** Results from regression models of frequency of maternal smoking during pregnancy and neonatal outcomes.

|  | Coefficient | p-value | Lower CI | Upper CI |
| --- | --- | --- | --- | --- |
| Birthweight (presented as kg from linear regression) |  |  |  |  |
| N = 1770 |  |  |  |  |
| Smoking during pregnancy: none | ref | ref | ref | ref |
| Yes, occasionally | -0.35 | <0.001 | -0.49 | -0.20 |
| Yes, most days | -0.12 | 0.524 | -0.50 | 0.25 |
| Yes, every day | -0.42 | <0.001 | -0.59 | -0.24 |
| Test for linear trend p-value |  |  |  | 0.001 |
| Low Birth Weight (presented as odds ratios from logistic regression) |  |  |  |  |
| N = 1770 |  |  |  |  |
| Smoking during pregnancy: none | ref | ref | ref | ref |
| Yes, occasionally | 3.08 | 0.012 | 1.28 | 7.40 |
| Yes, most days | 1.65 | 0.474 | 0.42 | 6.51 |
| Yes, every day | 6.04 | <0.001 | 2.95 | 12.36 |
| Test for linear trend p-value |  |  |  | <0.001 |
| Days in hospital (presented as days from negative binomial regression models) |  |  |  |  |
| N=1736 |  |  |  |  |
| Smoking during pregnancy: none | ref | ref | ref | ref |
| Yes, occasionally | 0.26 | 0.113 | -0.06 | 0.58 |
| Yes, most days | 0.14 | 0.538 | -0.32 | 0.60 |
| Yes, every day | 0.36 | 0.190 | -0.18 | 0.89 |
| Test for linear trend p-value |  |  |  | 0.268 |
| Being admitted to neonatal unit (presented as odds ratios from logistic regression) |  |  |  |  |
| N=1843 |  |  |  |  |
| Smoking during pregnancy: none | ref | ref | ref | ref |
| Yes, occasionally | 1.16 | 0.744 | 0.47 | 2.89 |
| Yes, most days | 0.82 | 0.765 | 0.22 | 3.07 |
| Yes, every day | 1.42 | 0.400 | 0.62 | 3.24 |
| Test for linear trend p-value |  |  |  | 0.64 |

Results from four separate models, each controlled for baby sex, nation of UK, Index of Multiple Deprivation (in five groups), maternal age and maternal ethnicity
CI = Confidence Interval
ref = reference category

**Appendix table 2:** Results from regression models of frequency of maternal vaping during pregnancy and neonatal outcomes.

|  | Coefficient | p-value | Lower CI | Upper CI |
| --- | --- | --- | --- | --- |
| Birthweight (presented as kg from linear regression) |  |  |  |  |
| N=1,627 |  |  |  |  |
| Vaping during pregnancy: none | ref | ref | ref | ref |
| Yes, occasionally | -0.22 | 0.041 | -0.42 | -0.01 |
| Yes, most days | 0.00 | 0.988 | -0.35 | 0.34 |
| Yes, every day | 0.03 | 0.83 | -0.22 | 0.27 |
| Test for linear trend p-value |  |  |  | 0.507 |
| Low Birth Weight (presented as odds ratios from logistic regression) |  |  |  |  |
| N=1,627 |  |  |  |  |
| Vaping during pregnancy: none | ref | ref | ref | ref |
| Yes, occasionally | 1.60 | 0.418 | 0.51 | 5.02 |
| Yes, most days | 1.08 | 0.945 | 0.13 | 8.74 |
| Yes, every day | 0.71 | 0.758 | 0.08 | 6.22 |
| Test for linear trend p-value |  |  |  | 0.69 |
| Days in hospital (presented as days from negative binomial regression models) |  |  |  |  |
| N=1,582 |  |  |  |  |
| Vaping during pregnancy: none | ref | ref | ref | ref |
| Yes, occasionally | 0.63 | 0.019 | 0.10 | 1.16 |
| Yes, most days | 0.14 | 0.534 | -0.29 | 0.56 |
| Yes, every day | -0.10 | 0.414 | -0.34 | 0.14 |
| Test for linear trend p-value |  |  |  | 0.148 |
| Being admitted to neonatal unit (presented as odds ratios from logistic regression) |  |  |  |  |
| N=1,658 |  |  |  |  |
| Vaping during pregnancy: none | ref | ref | ref | ref |
| Yes, occasionally | 1.40 | 0.487 | 0.54 | 3.66 |
| Yes, most days | 1.32 | 0.730 | 0.28 | 6.28 |
| Yes, every day* | * | * | * | * |
| Test for linear trend p-value |  |  |  | ** |

Results from four separate models, each controlled for baby sex, nation of UK, Index of Multiple Deprivation (in five groups), maternal age and maternal ethnicity
CI = Confidence Interval
\* Estimate not possible due to low numbers
\*\* Not estimable

**Appendix table 3.**
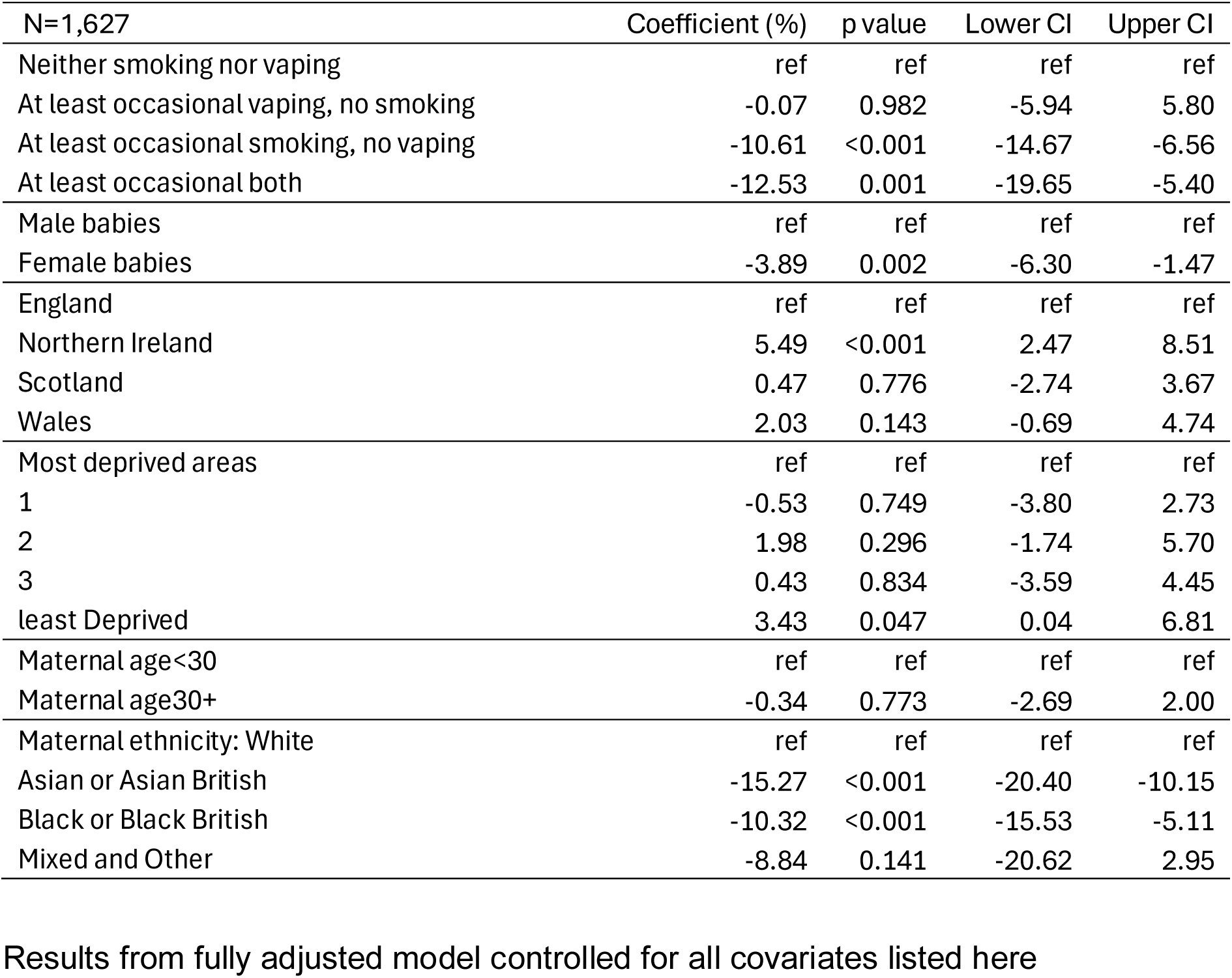
Results from linear regression model of maternal smoking/vaping during pregnancy and logged birthweight (results reported as percentages)

**Appendix table 4.** Analyses of outcomes using household income rather than Index of Multiple Deprivation.

|  | Coefficient | p-value | Lower CI | Upper CI |
| --- | --- | --- | --- | --- |
| Birthweight (presented as kg from linear regression) |  |  |  |  |
| N=1,504 |  |  |  |  |
| Neither smoking nor vaping | ref | ref | ref | ref |
| At least occasional vaping, no smoking | 0.01 | 0.922 | -0.14 | 0.16 |
| At least occasional smoking, no vaping | -0.32 | 0 | -0.45 | -0.18 |
| At least occasional both | -0.43 | 0 | -0.65 | -0.21 |
| Low Birth Weight (presented as odds ratios from logistic regression) |  |  |  |  |
| N=1,627 |  |  |  |  |
| Neither smoking nor vaping | ref | ref | ref | ref |
| At least occasional vaping, no smoking | 0.54 | 0.447 | 0.11 | 2.66 |
| At least occasional smoking, no vaping | 3.96 | 0 | 2.22 | 7.08 |
| At least occasional both | 3.67 | 0.019 | 1.24 | 10.83 |
| Days in hospital (presented as days from negative binomial regression models) |  |  |  |  |
| N=1,582 |  |  |  |  |
| Neither smoking nor vaping | ref | ref | ref | ref |
| At least occasional vaping, no smoking | -0.07 | 0.602 | -0.35 | 0.20 |
| At least occasional smoking, no vaping | -0.02 | 0.842 | -0.25 | 0.20 |
| At least occasional both | 0.88 | 0.001 | 0.39 | 1.38 |
| Being admitted to neonatal unit (presented as odds ratios from logistic regression) |  |  |  |  |
| N=1,680 |  |  |  |  |
| Neither smoking nor vaping | ref | ref | ref | ref |
| At least occasional vaping, no smoking | 0.65 | 0.462 | 0.21 | 2.04 |
| At least occasional smoking, no vaping | 1.02 | 0.943 | 0.53 | 1.96 |
| At least occasional both | 1.42 | 0.535 | 0.47 | 4.31 |

## REFERENCES

[1] Ko T-J, Tsai L-Y, Chu L-C, Yeh S-J, Leung C, Chen C-Y, et al. Parental Smoking During Pregnancy and Its Association with Low Birth Weight, Small for Gestational Age, and Preterm Birth Offspring: A Birth Cohort Study. Pediatr Neonatol 2014;55:20–7. 10.1016/j.pedneo.2013.05.005.

[2] Royal College of Physicians. Hiding in plain sight: Treating tobacco dependency in the NHS https://www.rcplondon.ac.uk/projects/outputs/hiding-plain-sight-treating-tobacco-dependency-nhs, 2018 n.d.

[3] Royal College of Physicians. Passive smoking and children. A report of the Tobacco Advisory Group of the Royal College of Physicians. 2010 n.d.

[4] Di H-K, Gan Y, Lu K, Wang C, Zhu Y, Meng X, et al. Maternal smoking status during pregnancy and low birth weight in offspring: systematic review and meta-analysis of 55 cohort studies published from 1986 to 2020. World J Pediatr 2022;18:176–85. 10.1007/s12519-021-00501-5.

[5] Lindson N, Butler AR, McRobbie H, Bullen C, Hajek P, Wu AD, et al. Electronic cigarettes for smoking cessation. Cochrane Database Syst Rev 2025;2025:CD010216. 10.1002/14651858.CD010216.pub9.

[6] Action on Smoking and Health. Use of e-cigarettes (vapes) among adults in Great Britain. https://ash.org.uk/resources/view/use-of-e-cigarettes-among-adults-in-great-britain 2025 n.d.

[7] Hopkinson NS. The prominence of e-cigarettes is a symptom of decades of failure to tackle smoking properly 2019. 10.1136/bmj.l647.

[8] Hopkinson NS. E-Cigarettes as a Smoking Cessation Aid - Toward Common Ground and a Rational Approach. Am J Respir Crit Care Med 2023;208:1007–9. 10.1164/rccm.202309-1623ed.

[9] Hopkinson NS, Vrinten C, Parnham JC, Radó MK, Filippidis F, Vamos EP, et al. Association of time spent on social media with youth cigarette smoking and e-cigarette use in the UK: a national longitudinal study 2024. 10.1136/thorax-2023-220569.

[10] Lange S, Probst C, Rehm J, Popova S. National, regional, and global prevalence of smoking during pregnancy in the general population: a systematic review and meta-analysis. Lancet Glob Health 2018;6:e769–76. 10.1016/S2214-109X(18)30223-7.

[11] McNeill A, Simonavičius E, Brose L, et al. Nicotine vaping in England: an evidence update including health risks and perceptions, 2022. https://assets.publishing.service.gov.uk/government/uploads/system/uploads/attachment_data/file/1107701/Nicotine-vaping-in-England-2022-report.pdf 2022 n.d.

[12] Bonello K, Figoni H, Blanchard E, Vignier N, Avenin G, Melchior M, et al. Prevalence of smoking during pregnancy and associated social inequalities in developed countries over the 1995–2020 period: A systematic review. Paediatr Perinat Epidemiol 2023;37:555–65. 10.1111/ppe.12989.

[13] Calder R, Gant E, Bauld L, McNeill A, Robson D, Brose LS. Vaping in Pregnancy: A Systematic Review. Nicotine Tob Res 2021;23:1451–8. 10.1093/ntr/ntab017.

[14] Ussher M, Fleming J, Brose L. Vaping during pregnancy: a systematic review of health outcomes. BMC Pregnancy Childbirth 2024;24:435. 10.1186/s12884-024-06633-6.

[15] Centre for Longitudinal Studies. Early Life Cohort Feasibility Study. . Available at https://cls.ucl.ac.uk/cls-studies/early-life-cohort-feasibility-study/ 2025 n.d.

[16] Centre for Longitudinal Studies (2025). Findings from the Early Life Cohort Feasibility Study survey component. Available at Findings-from-the-ELC-FS-survey-component.pdf n.d.

[17] Hopkinson NS, Bush A, Allinson JP, Faner R, Zar HJ, Agustí A. Early Life Exposures and the Development of Chronic Obstructive Pulmonary Disease across the Life Course. Am J Respir Crit Care Med 2024;210:572–80. 10.1164/rccm.202402-0432PP.

[18] U.S. Food and Drug Administration (FDA) Regulation of Electronic Nicotine Delivery Systems (ENDS): Background and Selected Policy Issues. 2025. Available at U.S. Food and Drug Administration (FDA) Regulation of Electronic Nicotine Delivery Systems (ENDS): Background and Selected Policy Issues | Congress.gov | Library of Congress n.d.

[19] Coleman T, Chamberlain C, Davey M-A, Cooper SE, Leonardi-Bee J. Pharmacological interventions for promoting smoking cessation during pregnancy - Coleman, T - 2015 | Cochrane Library n.d.

[20] Hajek P, Przulj D, Pesola F, Griffiths C, Walton R, McRobbie H, et al. Electronic cigarettes versus nicotine patches for smoking cessation in pregnancy: a randomized controlled trial. Nat Med 2022;28:958–64. 10.1038/s41591-022-01808-0.

[21] Damhuis SE, Ganzevoort W, Gordijn SJ. Abnormal Fetal Growth: Small for Gestational Age, Fetal Growth Restriction, Large for Gestational Age: Definitions and Epidemiology. Obstet Gynecol Clin North Am 2021;48:267–79. 10.1016/j.ogc.2021.02.002.

[22] Royal College of Obstetrics and Gynecology (2017) Small-for-Gestational-Age Fetus and a Growth Restricted Fetus, Investigation and Care (Green-top Guideline No. 31). Available at Small-for-Gestational-Age Fetus and a Growth Restricted Fetus, Investigation and Care (Green-top Guideline No. 31) | RCOG n.d.

[23] Simoncic V, Deguen S, Enaux C, Vandentorren S, Kihal-Talantikite W. A Comprehensive Review on Social Inequalities and Pregnancy Outcome—Identification of Relevant Pathways and Mechanisms. Int J Environ Res Public Health 2022;19. 10.3390/ijerph192416592.

[24] Jawad M, Cheeseman H, Brose LS. Waterpipe tobacco smoking prevalence among young people in Great Britain, 2013–2016. Eur J Public Health 2018;28:548–52. 10.1093/eurpub/ckx223.

